# Increased severity of influenza-associated hospitalizations in resource-limited settings: Results from the Global Influenza Hospital Surveillance Network (GIHSN)

**DOI:** 10.1101/2022.11.22.22282628

**Authors:** Lily E Cohen, Chelsea Hansen, Melissa K Andrew, Shelly A McNeil, Philippe Vanhems, Jan Kyncl, Javier Díez Domingo, Tao Zhang, Ghassan Dbaibo, Victor Alberto Laguna-Torres, Anca Draganescu, Elsa Baumeister, Doris Gomez, Sonia M Raboni, Heloisa I G Giamberardino, Marta C Nunes, Elena Burtseva, Anna Sominina, Snežana Medić, Daouda Coulibaly, Afif Ben Salah, Nancy A Otieno, Parvaiz A Koul, Serhat Unal, Mine Durusu Tanriover, Marie Mazur, Joseph Bresee, Cecile Viboud, Sandra S Chaves

**Affiliations:** Ready2Respond, Atlanta, Georgia, USA; The Task Force for Global Health, Atlanta, Georgia, USA; Icahn School of Medicine at Mount Sinai, New York, New York, USA; Division of International Epidemiology and Population Studies, Fogarty International Center, National Institutes of Health, Bethesda, Maryland, USA; Canadian Center for Vaccinology, IWK Health Centre and Nova Scotia Health, Dalhousie University, Halifax, Nova Scotia, Canada; Hôpital Edouard Herriot, Lyon, France; Department of Infectious Diseases Epidemiology, National Institute of Public Health, Prague, Czech Republic; Fundación para el Fomento de la Investigación Sanitaria y Biomédica de la Comunitat Valenciana (FISABIO-Public Health), Valencia, Spain; School of Public Health, Fudan University, Shanghai, China; Center for Infectious Diseases Research, American University of Beirut, Beirut, Lebanon; Clínica Internacional, Instituto de Medicina Tropical Universidad Nacional Mayor de San Marcos, Lima, Peru; National Institute for Infectious Diseases “Prof. Dr. Matei Bals”, Bucharest, Romania; Respiratory Virus Laboratory, Virology Department, INEI-ANLIS, Buenos Aires, Argentina; Grupo de Investigación UNIMOL, Facultad de Medicina, Universidad de Cartagena, Cartagena de Indias, Colombia; Virology Laboratory, Infectious Diseases Division, Universidade Federal do Paraná; Hospital Pequeno Principe, Curitiba, Paraná, Brazil; South African Medical Research Council, Vaccines and Infectious Diseases Analytics Research Unit, and Department of Science and Technology/National Research Foundation, South African Research Chair Initiative in Vaccine Preventable Diseases Faculty of Health Sciences, University of the Witwatersrand, Johannesburg, South Africa; Gamaleya Federal Research Center for Epidemiology and Microbiology, Ministry of Health of Russian Federation, Moscow, Russia; Smorodintsev Research Institute of Influenza, St Petersburg, Russia; Institute of Public Health of Vojvodina, Novi Sad; Faculty of Medicine, University of Novi Sad, Novi Sad, Serbia; National Institute of Public Hygiene (INHP), Abidjan, Côte d’Ivoire; Institut Pasteur de Tunis, Tunisia And Arabian Gulf University, Bahrain; Kenya Medical Research Institute (KEMRI), Nairobi, Kenya; Sheri Kashmir Institute of Medical Sciences, Srinagar, India; Department of Infectious Diseases and Clinical Microbiology, Hacettepe University School of Medicine, Ankara, Turkey; and Turkish Society of Internal Medicine, Turkey; Department of Internal Medicine, Hacettepe University School of Medicine, Ankara, Turkey; and Turkish Society of Internal Medicine, Turkey; Foundation for Influenza Epidemiology, Fondation de France, Paris, France

## Abstract

**Background:** Influenza disease data remain scarce in middle and lower-income countries. We used data from the Global Influenza Hospital Surveillance Network (GIHSN), a prospective multi-country surveillance system from 2012-2019, to assess differences in the epidemiology and severity of influenza hospitalizations by country income level.

**Methods:** We compiled individual-level data on acute respiratory hospitalizations, with standardized clinical reporting and testing for influenza. Adjusted odds ratios (aORs) for influenza-associated intensive care unit (ICU) admission and in-hospital death were estimated with multivariable logistic regression that included country income group (World Bank designation: high-income countries: HIC; upper middle-income countries: UMIC; lower middle-income countries: LMIC), age, sex, number of comorbidities, influenza subtype and lineage, and season as covariates.

**Findings:** From 73,121 patients hospitalized with respiratory illness in 22 countries, 15,660 were laboratory-confirmed for influenza. After adjustment for patient-level covariates, there was a two-fold increased risk of ICU admission for patients in UMIC (aOR 2.31; 95% confidence interval (CI) 1.85-2.88, p < 0.001), and a 5-fold increase in LMIC (aOR 5.35; 95% CI 3.98-7.17, p < 0.001), compared to HIC. The risk of in-hospital death in HIC and UMIC was comparable (UMIC: aOR 1.14; 95% 0.87-1.50; p > 0.05), though substantially lower than that in LMIC (aOR 5.05; 95% 3.61-7.03; p < 0.001 relative to HIC). A similar severity increase linked to country income was found in influenza-negative patients.

**Interpretation:** We found significant disparities in influenza severity among hospitalized patients in countries with limited resources, supporting global efforts to implement public health interventions.

**Funding:** The GIHSN is partially funded by the Foundation for Influenza Epidemiology (France). This analysis was funded by Ready2Respond under Wellcome Trust grant 224690/Z/21/Z.

**Research in Context:** *Evidence before this study:* In the past 35 years, fewer than 10% of peer-reviewed articles on influenza burden of disease have reported analyses from lower middle- or lower-income settings. Whereas the impact of influenza in upper middle- and high-income countries – regions where influenza seasonality is well-defined and where high numbers of influenza-related clinic visits, hospital admissions, and deaths are well-documented – has been clearly quantified, data scarcity has challenged our ability to ascertain influenza burden in resource-limited settings. As a result, policy decisions on vaccine use in lower-income countries have been made with limited data, slowing the development of influenza vaccine recommendations in these settings. In this study, we have conducted prospective influenza surveillance in the hospital setting in multiple countries to assess potential geographic differences in the severity of influenza admissions and have shown that influenza is a global concern, and report poorer clinical outcomes among patients admitted to hospitals in resource-limited settings. In these settings, it is especially important to consider the role of preventive measures, such as vaccines, in providing protection against severe disease.

*Added value of this study:* Since 2012, in collaboration with over 100 clinical sites worldwide, the Global Influenza Hospital Surveillance Network (GIHSN) has provided patient-level data on severe influenza-like illnesses based on a core protocol and consistent case definitions. To our knowledge, this is the first study to analyze multiple years of global, patient-level data generated by prospective, hospital-based surveillance across a large number of countries to investigate geographic differences in both influenza morbidity and mortality. Our study provides information on influenza burden in under-researched populations, particularly those in lower middle-income countries, and highlights the need for continued global collaboration and unified protocols to better understand the relationships between socio-economic development, healthcare, access to care, and influenza morbidity and mortality. After adjustment for differences in the characteristics of individual patients admitted to the hospital for influenza, we find an increased severity of disease in lower-income settings. In particular, the risk of ICU admissions increases two- and five-fold in upper middle- and lower-middle income countries, compared to high-income countries. The risk of in-hospital death is five-fold higher in lower-middle income countries, compared to more affluent countries.

*Implications of all the available evidence:* We find evidence of increased severity in influenza admissions in lower-income countries, which could point at structural differences in access to care between countries (patients arriving at the hospital later in the disease process) and/or differences in care once in the hospital. Understanding the mechanisms responsible for these disparities will be important to improve management of influenza, optimize vaccine allocation, and mitigate global disease burden. The Global Influenza Hospital Surveillance Network serves as an example of a collaborative platform that can be expanded and leveraged to address geographic differences in the epidemiology and severity of influenza, especially in lower and upper middle-income countries.

## Introduction

An estimated 3 to 5 million severe influenza cases occur worldwide each year, resulting in 290,000 to 650,000 influenza-related deaths.^1,2^ Patients with prior comorbidities are at high risk of severe influenza outcomes (e.g., hospital and/or intensive care unit admission, ventilation requirements, and death), but those who are otherwise healthy may also develop clinical complications.^3–5^ Given lack of routine testing for influenza, and the complex pathway by which influenza can exacerbate chronic conditions and lead to severe disease^6^, influenza burden estimates remain uncertain, especially in data-poor settings. Global estimates of influenza-related excess death require extrapolation of estimates from data-rich countries with established vital statistics systems to those without, with rare opportunities for direct validation in lower-income settings.^1,7^ And while most global studies have focused on influenza-associated respiratory disease deaths, few have studied hospital outcomes. Further, global estimation of influenza hospitalization rates remains challenging due to the heterogeneity of data collection methods and case definitions.^8^ Despite these limitations, regional and possibly income-based differences in rates of severe disease have been reported; for example, model estimates of influenza-associated respiratory mortality rates were highest in Sub-Saharan Africa and South-East Asia and lowest in Europe, the Western Pacific, and the Americas.^7^

The Global Influenza Hospitalization Surveillance Network (GIHSN) was established in 2011 to generate standardized patient-level data on severe disease across multiple countries and seasons and inform public health decision-making.^9^ The GIHSN uses a prospective, active, hospital-based surveillance approach to collect epidemiological and virological data from admitted patients meeting eligibility criteria and a standard definition of influenza-like-illness (ILI).^10,11^ In this paper, we explore geographic differences in the global epidemiology and severity of influenza based on the GISHN data and compare clinical outcomes in high- (HIC), upper middle-(UMIC), and lower middle-income (LMIC) countries.

## Methods

### Overview of surveillance network

We analyzed GIHSN data from 2012 through 2019. The network has expanded from 11 hospitals coordinated by four partner sites in three countries in its inception in 2011 to 116 hospitals coordinated by 24 partner sites in 22 countries (Figure 1; supplementary Table S1, Figure S1 shows the contribution and patient population of each site).

**Figure 1:**
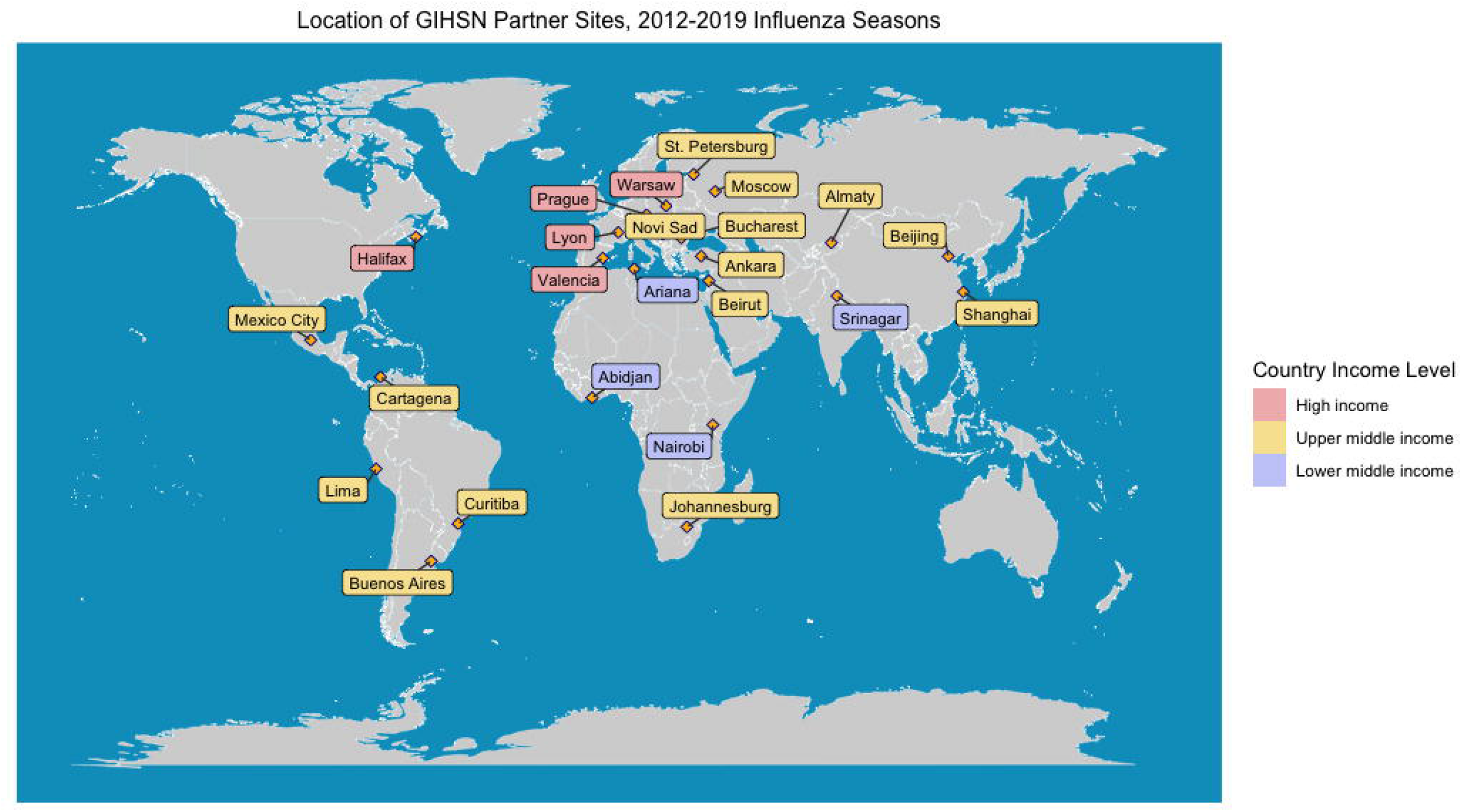
Geographic distribution of sites collaborating in the Global Influenza Hospital Surveillance Network (GIHSN), from 2012 through 2019 seasons

The GIHSN uses a core protocol to identify and recruit eligible patients (supplementary Figure S2). Patients are considered eligible for the study if they are admitted through the emergency department or into the inpatient ward of a participating hospital for acute (onset within seven days prior to hospital admission) respiratory illness defined as a combination of at least one systemic symptom (fever or feverishness, headache, myalgia, or malaise); and at least one respiratory symptom (cough, sore throat, shortness of breath, or nasal congestion^11^). Patients are enrolled within 72 hours of hospital admission and need to be hospitalized for at least one night. Patients are excluded from the study if they are unable to communicate or consent to participation. Respiratory specimens are collected from enrolled patients using nasal-pharyngeal and/or oral-pharyngeal swabs that are tested for influenza by reverse transcription polymerase chain reaction (RT-PCR). Influenza-positive samples are sequenced to identify specific influenza A subtypes or B lineages. Clinical and demographic information is collected for each enrolled patient with a questionnaire at admission and through medical chart review at discharge. Clinical data collected include duration of hospital stay, presence of comorbidities, history of influenza vaccination, admission to intensive care unit, use of mechanical ventilation and date of discharge or death. Comorbidities include cardiovascular disease, chronic obstructive pulmonary disease, diabetes, asthma, neuromuscular disease, cirrhosis, renal disease, and immunosuppression (comprising autoimmunity, neoplasm, rheumatologic disease, and other immunosuppressive conditions such as HIV/AIDS). All data are anonymized and centralized for analysis.

### Statistical analyses

#### Descriptive analyses

We summarized patients’ demographic and clinical characteristics, such as age (<5 years, 5-64 years, and ≥65 years), sex, number and type of comorbidities, smoking status, and whether they were treated with antivirals in the hospital. We assessed patients with and without laboratory confirmed influenza separately. We also described the characteristics of patients by country income level (Gross National Income (GNI) per capita, World Bank^12^), hemisphere, influenza type, subtype or lineage, and case severity (defined as ICU admission and death). More details are provided in supplementary material Table S2.

#### Risk factor analyses

Multivariable logistic regression models were used to estimate adjusted odds ratios (aORs) for severe clinical outcomes: specifically, intensive care unit (ICU) admission and in-hospital death among influenza-positive patients. Covariates considered included country income level, sex, age category, number of comorbidities, season, and influenza subtype or lineage. For all covariates, differences were considered significant at *p* < 0.05. Akaike information criteria (AIC) was used to compare models containing different subsets of covariates. Data analysis was performed using R version 4.1.2 software.

#### Sensitivity Analysis

To assess whether the risk factors for severe outcomes evidenced in the influenza data were specific to influenza or common to other respiratory illnesses, we ran the same regression models for ICU admission and death in the influenza-negative population, using identical covariates as in the main analysis save influenza subtype or lineage.

## Results

### Patient population, and distribution of influenza strains and seasonality

From 2012-2019, a total of 73,121 hospitalized patients met surveillance inclusion criteria (supplementary Table S3), of whom 15,560 (21.3%) were influenza-positive. The majority of influenza positive samples (69.2%) were from UMIC, while HIC contributed 26.6% and LMIC contributed 4.2% (supplementary Figure S3).

Overall, among the 15,560 influenza-positive patients, 72.4% had influenza A, and 27.6% influenza B. Influenza A/H3N2 was the predominant subtype (n=5,522; 35.5%), followed by A/H1N1pdm09 (n=4917; 31.6%). Notably, as country income level declined, the share of A/H1N1pdm09 increased, with A/H3N2 and A/H1N1pdm09 identified in almost the same number of patients in LMIC. Of the type B influenza strains, B/Yamagata was the most common in all income groups (n=2,226; 51.6%, p < 0.001). B/Victoria comprised a larger proportion of the influenza-positive patients in UMIC and LMIC than it did in HIC.

Influenza positivity varied between years (median of 12.5% in LMIC across all seasons, 24.6% in UMIC across all seasons, and 14.8% in HIC across all seasons) (Figure 2). In HIC (all countries were in the Northern Hemisphere), median percent positivity was highest from December to March (31.8% - 50%) and lowest in September and October (0.46% and 0%, respectively). No samples were collected from HIC in July or August (considered off-season). In UMIC, the peak in median percent positivity differed by hemisphere. For the 9 countries UMIC in the Northern Hemisphere, median percent positivity was highest from December to March (25.5% - 31.9%) and lowest in May and October (2.0% and 0.36%, respectively). For the 4 UMIC countries in Southern Hemisphere, median percent positivity was highest from June to September (5.5% - 9.6%) and lowest from December to February (0%). Overall, samples from UMIC in the Southern Hemisphere had the lowest median percent positivity of all regions. Median percent positivity in LMIC, half of which were equatorial, was highest in February, June, and November (14.7% - 19.1%) and lowest in May and October (2.4% and 2.5%, respectively). The 2017/2018 season had the highest percent positivity, particularly for countries in the Northern Hemisphere.

**Figure 2:**
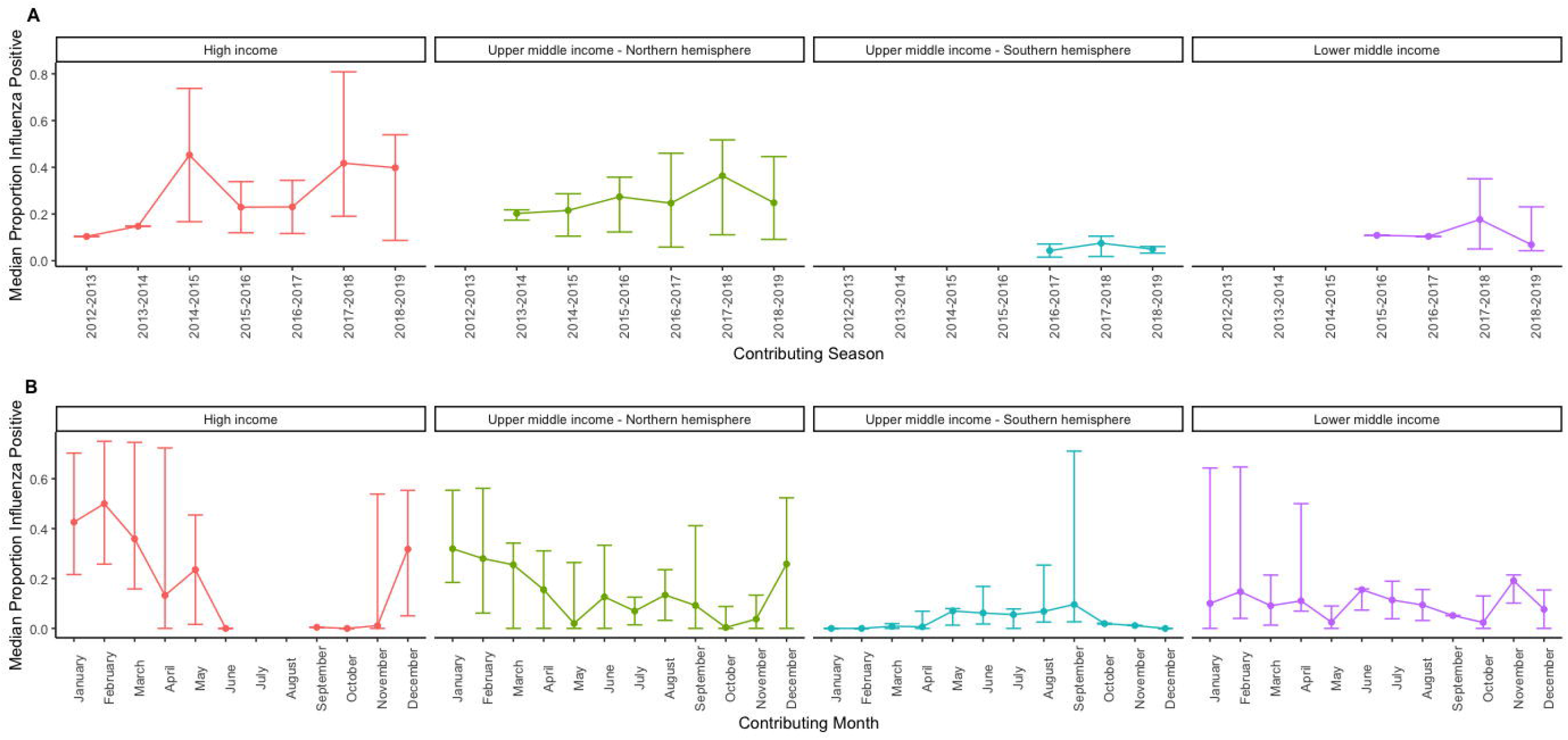
A) Median proportion and range of the percentage positivity for influenza by season and country income level; B) Median proportion and range of the percentage positivity for influenza by month and country income level, GIHSN 2012-2019

### Demographic and clinical characteristics of laboratory-confirmed influenza hospitalizations

Of the 15,660 influenza-positive patients, 28.7% were <5 years, 46.9% were 5-64 years, and 24.4% were ≥65 years (Table 1). The gender balance was similar across income groups. Overall, 48.4% of influenza-positive patients did not have any comorbidities, 29.9% had one, and 21.7% had two or more (Figure 3). The most common comorbidities across income levels were cardiovascular disease (21.0%) and immunosuppression due to rheumatologic or autoimmune disease, or neoplasm (27.4%). Presence of cardiovascular disease was more frequent in HIC (44.1%), as compared to UMIC (11.6%) and LMIC (29.7%), while presence of immunosuppressive conditions was more common in LMIC (51.5%), as compared to HIC (26.5%) and UMIC (26.3%). 58.0% of influenza-positive patients were non-smokers (51.4% in HIC, 60.1% in UMIC, 64.7% in LMIC; p < 0.001) (Table 1).

**Table 1:**
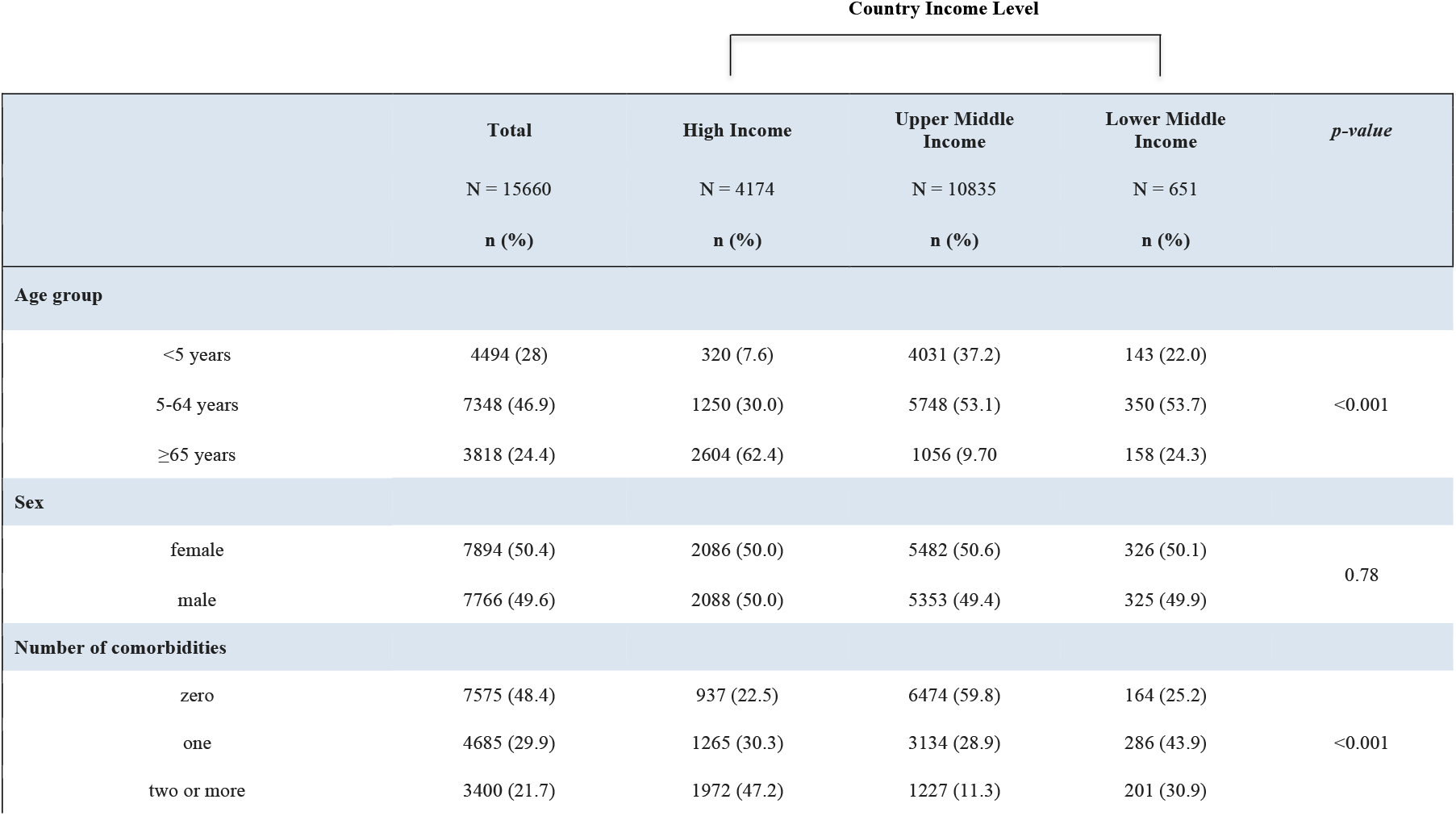
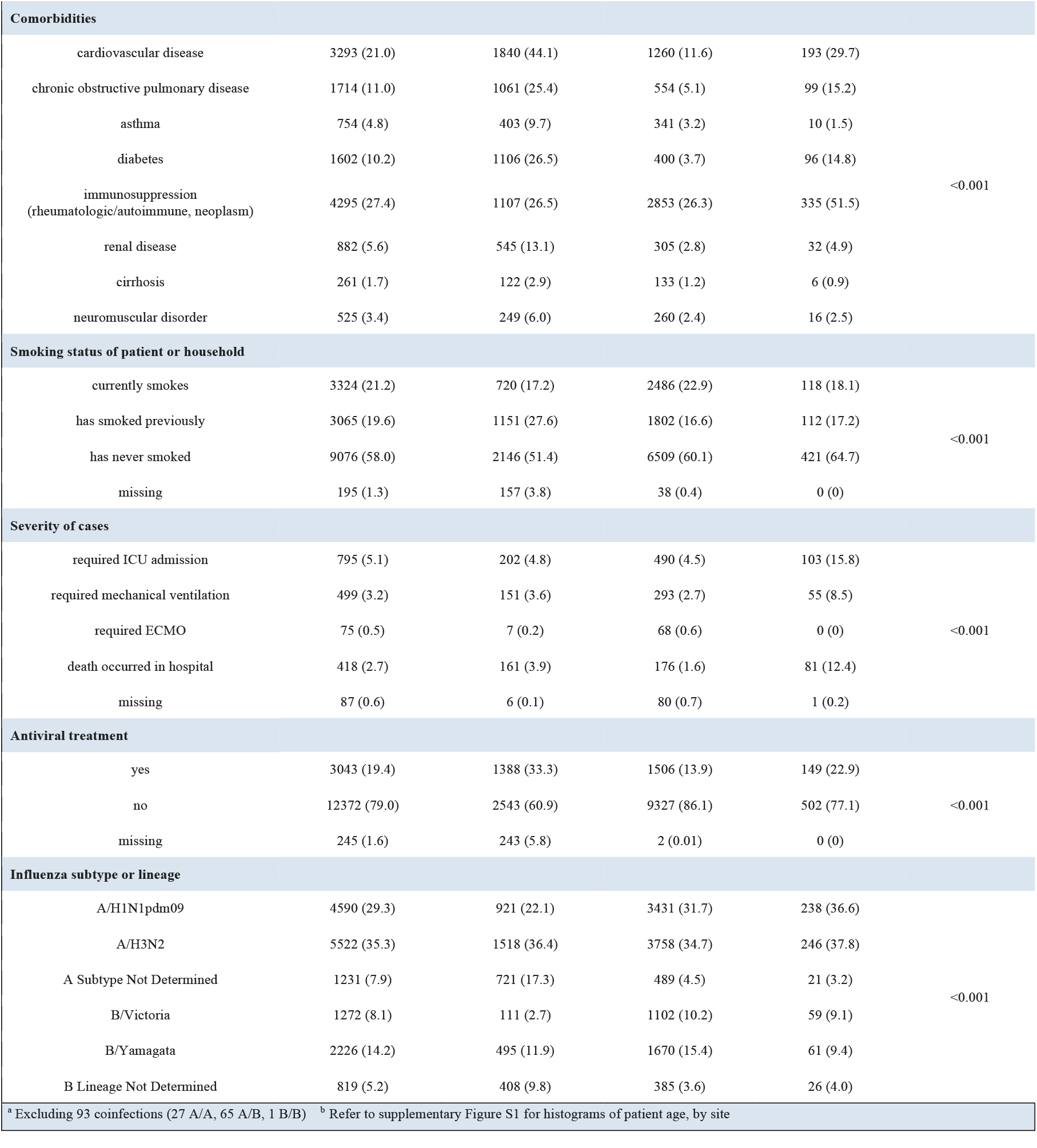
Demographic characteristics of patients hospitalized with laboratory-confirmed influenza by country income level, GIHSN 2012-2019

**Figure 3:**
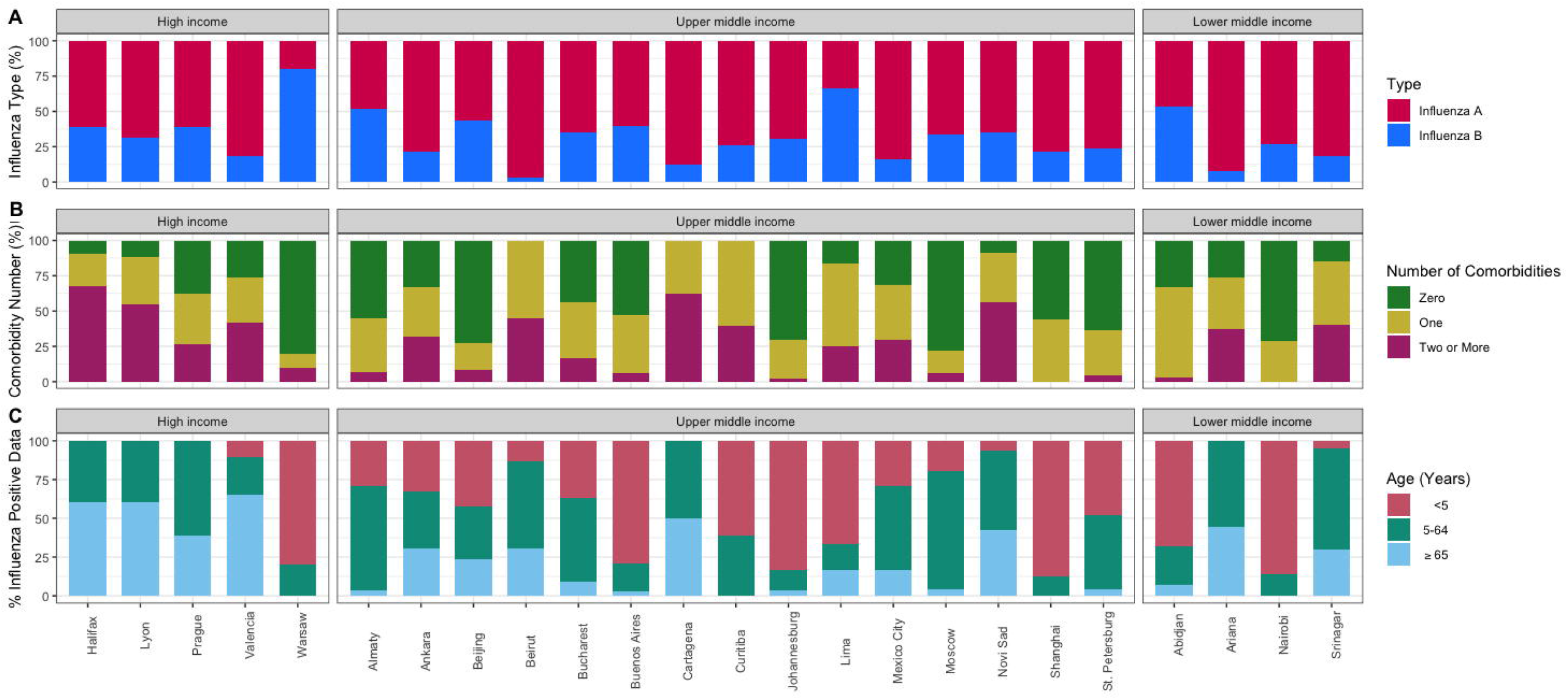
A) Distribution of influenza-positive hospitalizations by influenza type, site, and country income level; B) Distribution of influenza-positive hospitalizations by number of patient comorbidities, site, and country income level; C) Distribution of influenza-positive hospitalizations by age group, site, and country income level, GIHSN 2012-2019

In terms of clinical course, 5.1% of influenza-positive patients were admitted to the ICU, 3.2% received mechanical ventilation, 0.5% were placed on extracorporeal membrane oxygenation (ECMO), and 2.7% died while in hospital. Only 19.4% of influenza-positive patients were treated with an antiviral: 33.3% in HIC, 13.9% in UMIC, and 22.9% in LMIC (p < 0.001).

### Risk of influenza-associated ICU admission and in-hospital death

In total, 2.8% of patients <5 years were admitted to the ICU and 0.27% died in-hospital. Among patients 5-64 years, 5.2% were admitted to the ICU and 2.1% died in hospital, while 7.5% of patients ≥65 years were admitted to the ICU and 6.6% died in-hospital. Figure 4 shows differences in severity by income brackets, influenza type, and age group.

**Figure 4:**
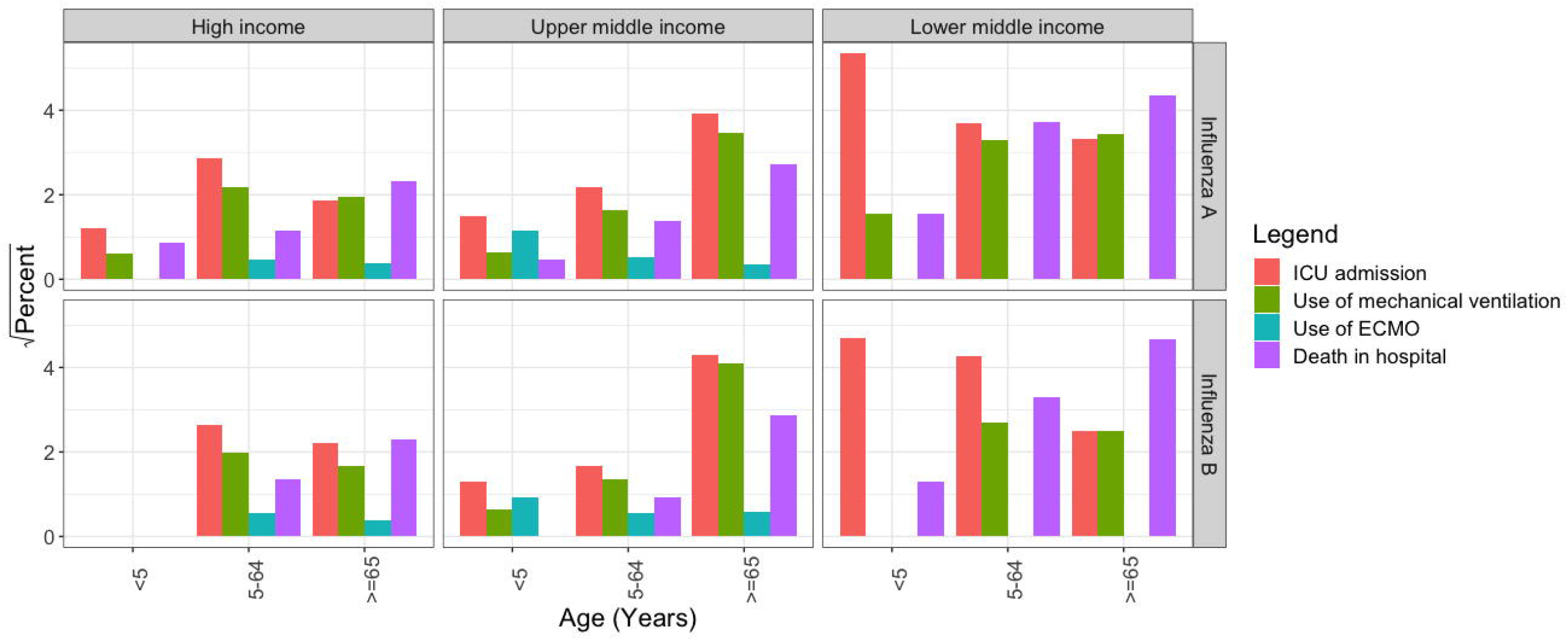
Severity among patients hospitalized with laboratory-confirmed influenza, by age group, influenza type and country income level, GIHSN 2012-2019

In the multivariable logistic regression, we found a two-fold increased risk of ICU admission for patients in UMIC (aOR 2.31; 95% confidence interval (CI) 1.85-2.88, p < 0.001), and 5-fold increase in LMIC (aOR 5.35; 95% CI 3.98-7.17, p < 0.001), compared to HIC (Figure 5). The risk of in-hospital death was higher in LMIC (aOR 5.05; 95% 3.61-7.03; p < 0.001) relative to HIC, but there was no statistically significant difference between UMIC and HIC. The presence of two or more comorbidities significantly increased the odds of ICU and death (aOR 5.71, 95% 4.40-7.43; p < 0.001 and aOR 3.12, 95% 2.16-4.56; p < 0.001, respectively). Female sex reduced the odds of ICU admission (aOR 0.80; 95% CI 0.69-0.93; p < 0.01) and in-hospital death (aOR 0.74; 95% CI 0.60-0.90; p < 0.01).

**Figure 5:**
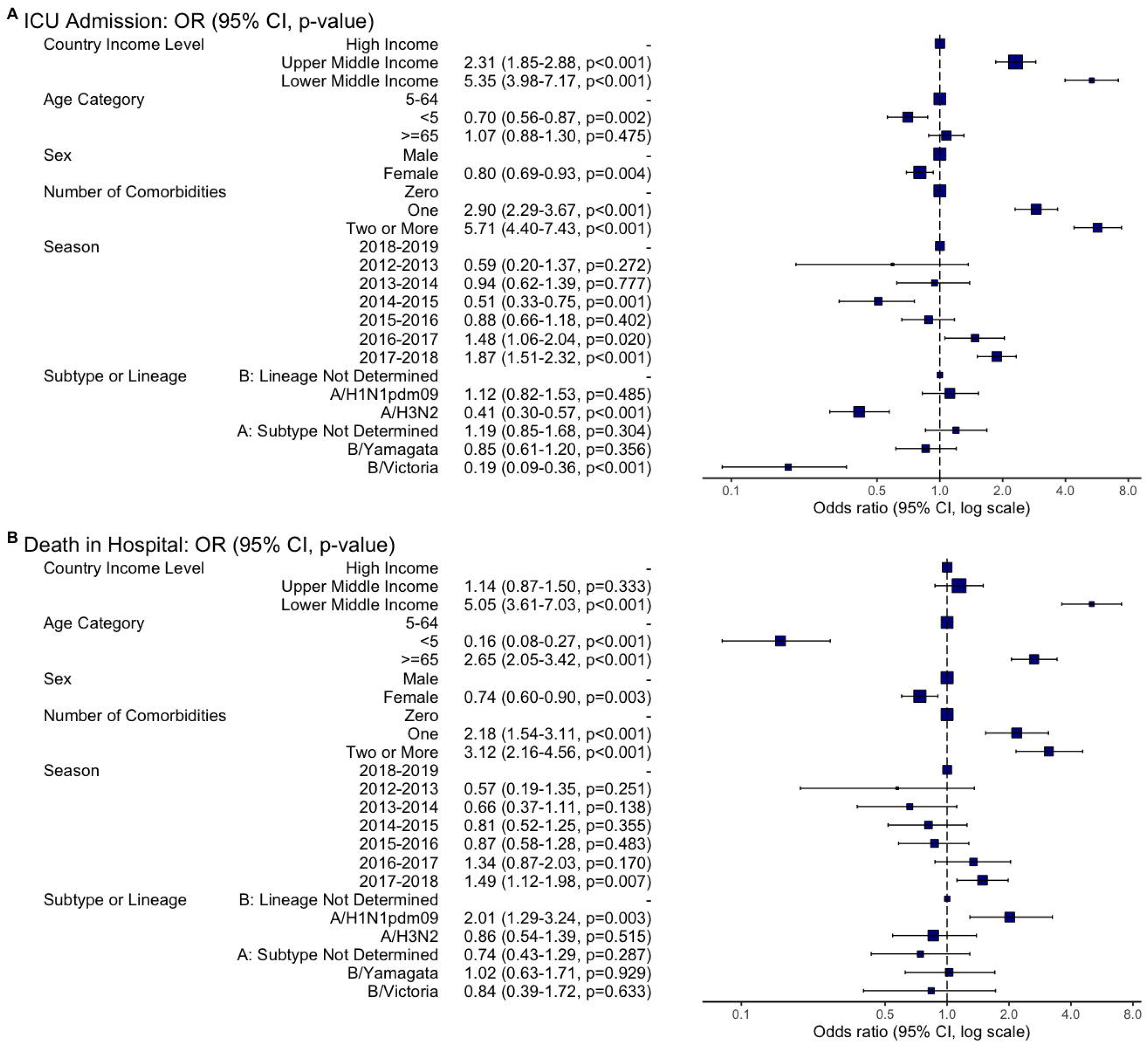
Risk factors for (A) intensive care unit admission and (B) in-hospital death among patients hospitalized with laboratory-confirmed influenza

Compared to patients 5-64 years, patients ≥65 years had higher odds of death but not ICU admission (aOR for death 2.65; 95% CI 2.05-3.42; p < 0.001). Patients <5 years had lower odds for both ICU admission (aOR 0.70; 95% CI 0.56-0.87; p < 0.01) and in-hospital death (aOR 0.16; 95% CI 0.08-0.27; p < 0.01). Being admitted during the 2017-2018 season increased the risk of ICU admission (aOR 1.87; 95% CI 1.51-2.32; p < 0.001) and in-hospital death (aOR 1.49; 95% CI 1.12-1.98; p < 0.01) approximately three-fold. Infection with A/H1N1pdm09 was an important independent predictor of death (aOR 2.01; 95% CI 1.29-3.24; p < 0.01) while A/H3N2 reduced the odds of ICU admission (aOR 0.41; 95% CI 0.30-0.57; p < 0.001). Pairwise comparison of odds ratios using Tukey’s honest significance test showed a statistically significant difference between A/H3N2 and A/H1N1pdm09, with the latter causing more severe illness, both when adjusting for age as a covariate (ICU: p < 0.001, death in hospital: p < 0.001) and when the dataset was restricted to only influenza-positive patients ≥65 years (ICU: p < 0.001, death in hospital: p < 0.05) (supplementary Table S4).

### Sensitivity analysis

As a sensitivity analysis, we ran the same regression model on patients hospitalized with severe acute respiratory illness but who were influenza-negative. Overall, many of the demographic and seasonal factors affecting influenza severity also affected influenza-negative patients, including increased risk of ICU admission and in-hospital death during the 2017/2018 season (supplementary Figure S4). However, the 2017/2018 season was not significant as a predictor of clinical severity when restricting analysis to HIC. The most notable difference was that influenza-negative children had a slightly higher odds ratio for ICU admission (aOR 1.13; 95% CI 1.02-1.25; p < 0.05) compared to the other age groups, contrasting with young age being a protective factor for ICU admission among influenza-positive patients.

## Discussion

In this study, we identify predictors of severity among patients presenting to hospitals in high-, upper middle-, and lower middle-income settings. We find that even when adjusting for age, sex, prevalence of comorbidities, and season, patients hospitalized with influenza were more likely to be admitted to the ICU or to die if they were in LMIC or UMIC compared to HIC. Independent predictors of severity also included older age (≥65 years), male sex, and having multiple comorbidities. Infection with influenza A/H1N1pdm09 also led to more severe outcomes, compared to other influenza viruses, even in patients ≥65 years. Though influenza is a significant global health issue, few studies have focused on the comparative epidemiology and severity of hospitalized patients by country income levels. As patient outcomes are driven in part by demographic and clinical factors, it is important to adjust for individual level characteristics before assessing broad geographic differences in influenza severity.

We detected seasonal patterns of influenza circulation broadly consistent with other studies, with winter peaks in the Northern and Southern Hemispheres and year-round activity in equatorial countries.^14^ Although A/H3N2 was the most frequently detected strain in all countries, A/H1N1pdm09 was more common in hospitalizations from LMIC. This could reflect differences in country demographics as A/H1N1pdm09 has been found to be more severe in children and young adults,^15^ who may have been over-represented in LMIC reporting, or the fact that LMIC contributed data in fewer seasons that happened to be A/H1N1pdm-dominant by chance. Additionally, we find increased severity of A/H1N1pdm09, even in patients ≥65 years. Prior analyses have reported clinical protection against severe A/H1N1pdm09 in older individuals from previous exposure to similar viruses^16^; here, it is likely that only a small percentage of enrolled patients would have been born early enough to enjoy such protection (i.e., born before 1950s).

Overall, influenza-related outcomes were generally worse in LMIC and UMIC, after adjusting for patient characteristics, including number of comorbidities. Several factors may be contributing to this association including the type of underlying medical conditions. Overall, immunosuppression was less prevalent in HIC, while immunocompromised individuals are at higher risk of severe influenza-related outcomes including death.^17^ Moreover, conditions such as malaria and malnutrition are likely more prevalent in LMIC and could have affected clinical prognosis, but were not recorded. Further, we could not adjust for vaccination in our models due to a large amount of missing information (70%). Influenza vaccination is generally less common in LMIC compared to HIC.^18-20^ and country-level recommendations differed substantially between participating sites.^21^ Moreover, differences in standards of care and/or lack of access to antivirals in LMIC are also a possible explanation: compared to HIC, fewer influenza-positive patients in LMIC and UMIC received antiviral therapy for influenza based on our data.

Severity disparities were particularly pronounced for ICU admissions; 23.7% of patients admitted to ICUs died in LMIC hospitals, whereas the proportion was 13.4% in HIC (data not shown). Intensive care units in LMIC may function differently than in high-income countries; the COVID-19 pandemic showed that some ICUs in LMIC had unclear thresholds for initiating mechanical ventilation and limited access to and/or usage of ECMO machines^22^ and decreased bed capacity.^23^ Given the lower capacity of ICUs in LMIC, it is worth noting that a large percentage of influenza-positive patients in our dataset had severe enough illness to meet the threshold for ICU admission (15.8%). It has been hypothesized that delayed care-seeking in LMIC could contribute to the increased severity of incoming patients.^24^

Interestingly, many of the geographic disparities and risk factors identified for influenza severity were present among influenza-negative patients. Patients in UMIC continued to have an increased risk of ICU admission with non-influenza respiratory illness (aOR 2.39; 95% 2.11-2.70; p < 0.001, relative to HIC). Risk factors, such as multiple comorbidities and male sex increased the odds of both ICU admission and in-hospital death, irrespective of the presence of influenza. Prior work has shown that comorbidities and sex affect severity of multiple respiratory conditions.^25,26^ Surprisingly, however, we found that the 2017-2018 season was more severe for both influenza and non-influenza patients. This season was a notably severe influenza A/H3N2 season in multiple countries,^27,28^ however it remains unclear why this season would be associated with increased severity of other respiratory pathogens. One hypothesis is the possible misclassification of patients due to delay in seeking care, i.e., some influenza-negative patients could actually have been positive if tested earlier on in the course of illness. An alternative, and less plausible hypothesis, is the role of co-infections, in that an undetected pathogen co-circulated with influenza during the 2017-2018 causing also severe respiratory disease. Notably, the effect of the 2017-2018 season disappeared when restricting the analysis to high-income countries (Figure S4) and there was also variability in the contribution of different sites each season. Sensitivity analyses excluding one site at a time suggested that overrepresentation of more severe sites, and underrepresentation of milder sites, affected the estimated risk in 2017-2018, although it did not fully explain it. Overall, given the heterogeneity in participating sites, their patients, and variation in pathogen circulation in each season, the seasonal term in our model likely captures multiple unexplained factors.

A notable difference between our analysis of influenza and non-influenza patients was that young age (<5 years) was protective against ICU admission in the context of influenza infection but was a moderate risk factor when influenza was absent. This likely speaks to the well-known contribution of respiratory syncytial virus (RSV) and metapneumovirus to severe disease among young children.^29-31^

Many studies, including past publications from the GIHSN, have called for increased distribution and uptake of influenza vaccines in low-, lower-, and upper middle-income countries.^21,33^ Yet, many low- and lower middle-income countries do not have national influenza vaccination policies.^33^ Although we were not able to investigate the impact of influenza vaccination programs on disease severity due to limited reporting and/or use of the influenza vaccine in most GIHSN countries, our main result – that influenza severity among hospitalized patients is increased in resource-limited settings –provides important support for interventions to mitigate influenza in LMIC and UMIC. Nevertheless, the benefits of influenza vaccination in each country must be considered alongside interventions for other diseases, requiring local data on disease burden and cost-effectiveness analyses. This is an important gap for future research, especially as new or improved interventions are becoming available for other respiratory pathogens of importance, such as SARS-CoV-2 and RSV.

Our data reflect influenza and non-influenza hospitalization patterns in the pre-COVID19 pandemic period. We excluded the 2019-2022 influenza seasons in our analyses due to multiple reports of depressed circulation of influenza and other endemic viruses during COVID-19.^34^ COVID-19 is now being monitored by the GIHSN, which could warrant further analyses.^35^ Further, assessing changes in the severity and epidemiology of influenza in the post-pandemic period will be important, as out-of-season activity and age shifts have been reported for multiple endemic pathogens.^36,37^

There are limitations to this study. We did not have a large representation of lower-income sites. Because the GIHSN has only been in operation for the past decade, additional collaborating sites are still being recruited, particularly in lower-income regions. Currently, the majority of sites fall in UMIC, and lower-income sites only joined the network surveillance recently. Further, the definition of intensive care units and/or intensive care may differ between sites and countries. Moreover, we did not have sufficient individual-level data on vaccination.^38^ Additionally, we lack a denominator to estimate influenza morbidity and mortality rates. Relatedly, we missed patients who are too ill to travel to the hospital,^39-41^ as well as those who die after hospital discharge.^42^

In conclusion, our study highlights the value of unified surveillance protocols to better understand the relationships between socio-economic development, healthcare, and influenza morbidity and mortality. We find that hospitalizations for influenza and other respiratory pathogens are more severe in lower-income settings, compared to more affluent countries. Our results confirm that influenza is a significant global public health concern, supporting global efforts to increase surveillance and mitigation efforts. Further research is warranted to clarify the mechanisms driving global disparities in the severity of respiratory diseases.

## Data Availability

Anonymized data used for this analysis, along with a data dictionary, are available upon request made to. The use of data depends on the approval of an analytical proposal by the Independent Scientific Committee. Investigators from participant sites are informed up front for any planned data analysis and they have the possibility to opt-out.

## Contributors

LEC, CH, CV and SSC conceived and designed the study. LEC and CH analyzed the data. LEC wrote the first draft of the manuscript, with substantial participation of CH, CV and SSC. All authors contributed, reviewed, and approved this Article. LEC and CH accessed and verified the data. All authors had final responsibility for the decision to submit for publication.

## Acknowledgments

We would like to thank Catherine Commaille-Chapus and Camile Hunsinger from Impact Healthcare for their assistance in network coordination and operational implementation, including data collection, verification, and hosting.

## Disclaimer

The findings and conclusions in this report are those of the authors and do not necessarily represent the official position of the U.S. National Institutes of Health or Department of Health and Human Services.

