## Supplementary Material for "Increased severity of influenza-associated hospitalizations in resource-limited settings: Results from the Global Influenza Hospital Surveillance Network (GIHSN)"

### Supplementary Information

Table S1: Characteristics of included sites, 2012-2019 influenza seasons

| Coordinating site | Included patient specimens | Number of contributing seasons | Country income level[a] | Specimen distribution (+/- flu) | Distribution: number of comorbidities |
|---|---|---|---|---|---|
| Canada (Halifax) | 1,351 | 2 | High income | | |
| France (Lyon) | 211 | 2 | High income | | |
| Czech Republic (Prague) | 461 | 4 | High income | | |
| Spain (Valencia) | 21,304 | 7 | High income | | |
| Poland (Warsaw) | 22 | 1 | High income | | |
| Kazakhstan (Almaty) | 259 | 1 | Upper middle income | | |
| China (Beijing) | 4,656 | 3 | Upper middle income | | |
| Lebanon (Beirut) | 522 | 1 | Upper middle income | | |
| Romania (Bucharest) | 1,822 | 3 | Upper middle income | | |
| Argentina (Buenos Aires) | 902 | 2 | Upper middle income | | |
| Colombia (Cartagena) | 42 | 1 | Upper middle income | | |
| Brazil (Curitiba) | 378 | 1 | Upper middle income | | |
| Turkey (Ankara) | 2,373 | 4 | Upper middle income | | |
| South Africa (Johannesburg) | 4,705 | 3 | Upper middle income[b] | | |
| Peru (Lima) | 530 | 3 | Upper middle income | | |
| Mexico (Mexico City) | 1,971 | 4 | Upper middle income | | |
| Russian Federation (Moscow) | 8,549 | 6 | Upper middle income | | |
| Serbia (Novi Sad) | 1,386 | 2 | Upper middle income | | |
| China (Shanghai) | 2,764 | 3 | Upper middle income | | |
| Russian Federation (St. Petersburg) | 14,184 | 6 | Upper middle income | | |
| Côte d'Ivoire (Abidjan) | 983 | 2 | Lower middle income | | |
| Tunisia (Ariana) | 77 | 1 | Lower middle income | | |
| Kenya (Nairobi) | 1,002 | 2 | Lower middle income | | |
| India (Srinagar) | 2,667 | 4 | Lower middle income | | |
| All sites | 73,121 | | | | |
| [a]According to World Bank GNI per capita, calendar year 2019 | | | | Flu A Flu B Flu - | 0 1 2+ |
| [b]Romania designated upper middle income in 2 out of 3 years of contribution to network | | | | | |

Figure S1: Age distribution of patients enrolled by site, and whether they were influenza-laboratory confirmed, GIHSN, 2012-2019

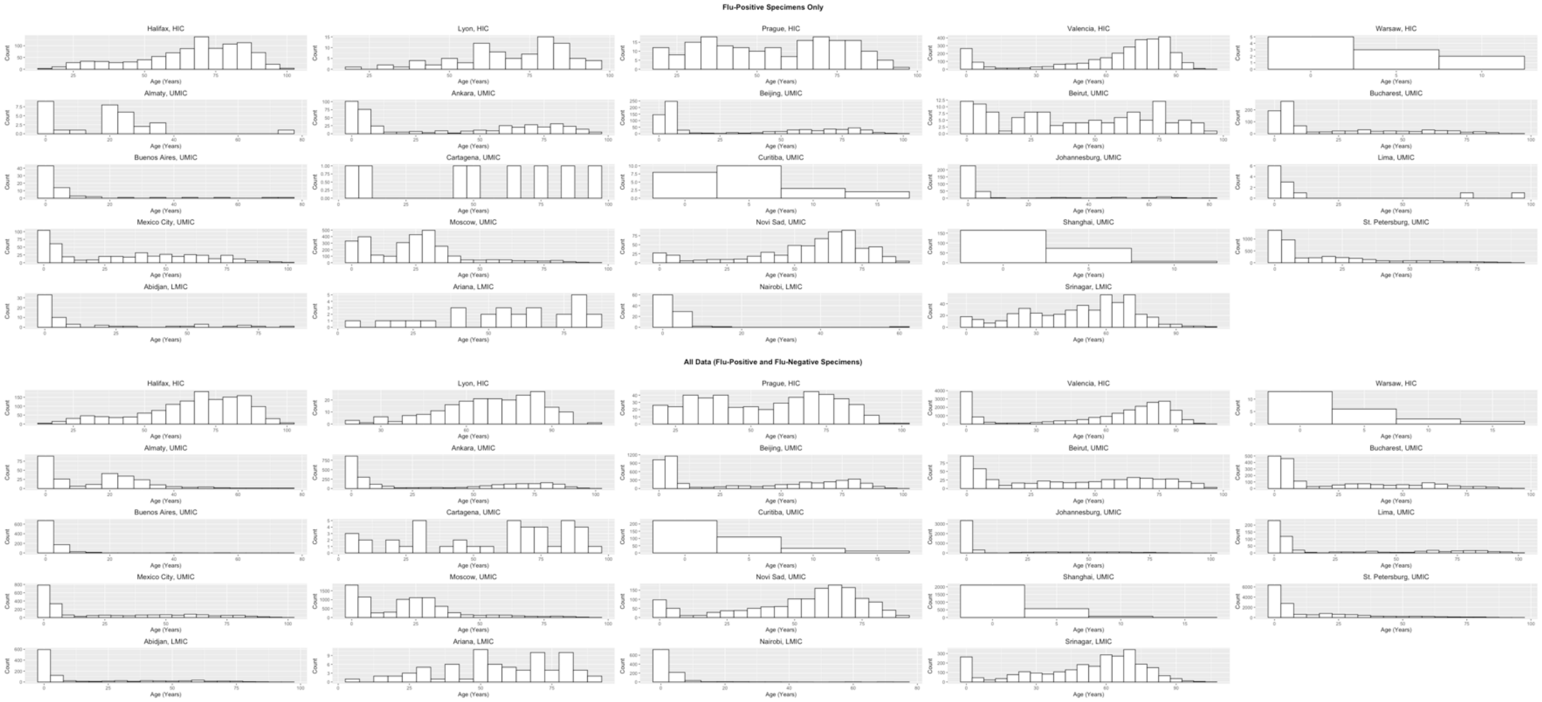

Table S2: Collation of dataset for analysis

| |
|---|
| We removed 86 specimens serving as internal controls for the coordinating sites and 1531 specimens missing patient age. We then removed 54,954 specimens for which questionnaires were partially or fully completed but which did not meet GIHSN criteria for inclusion (i.e., patients were recently discharged, were institutionalized, or were not residents of the hospital catchment area). Finally, we removed 279 specimens with inconsistent subtype or lineage data (e.g., a "yes" for A/H1N1pdm09 and a "yes" for unable to be subtyped) and 93 influenza-positive specimens with coinfections of more than one influenza strain.<br><br>The data used in the regression analyses of the influenza-positive data excludes 88 influenza-positive specimens for which ICU admission information is unavailable (ICU model) and 91 influenza-positive specimens for which it is unknown whether the patient died in the hospital (in-hospital death model). The data used in the regression analysis of influenza-negative specimens excludes 352 influenza-negative specimens for which ICU admission information is unavailable (ICU model), 321 influenza-negative specimens for which it is unknown whether the patient died in the hospital (in-hospital death model), and 257 influenza-negative specimens for which comorbidity information is not recorded (both models). |

Figure S2: GIHSN data collection protocol

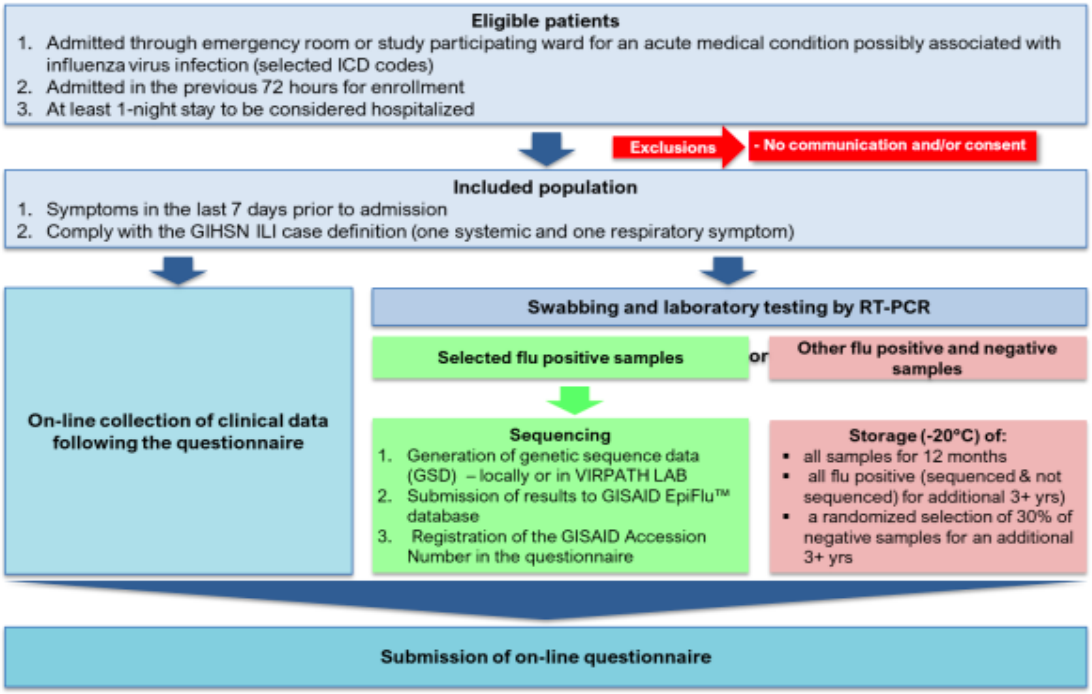

Table S3: Demographic characteristics of all patients hospitalized with severe acute respiratory illness by country income level, GIHSN 2012-2019

| | Total | Country Income Level | | | *p-value* |
|---|---|---|---|---|---|
| | | **High Income** | **Upper Middle Income** | **Lower Middle Income** | |
| | N = 73121 | N = 23349 | N = 45043 | N = 4729 | |
| | **n (%)** | **n (%)** | **n (%)** | **n (%)** | |
| **Age group** | | | | | |
| <5 years | 29450 (40.3) | 4460 (19.1) | 23149 (51.4) | 1841 (38.9) | |
| 5-64 years | 25191 (34.5) | 5579 (23.9) | 17845 (39.6) | 1767 (37.4) | <0.001 |
| ≥65 years | 18480 (25.3) | 13310 (57.0) | 4049 (9.0) | 1121 (23.7) | |
| **Sex** | | | | | |
| female | 34161 (46.7) | 10833 (46.4) | 21075 (46.8) | 2253 (47.6) | 0.26 |
| male | 38960 (53.3) | 12516 (53.6) | 23968 (53.2) | 2476 (52.4) | |
| **Number of comorbidities** | | | | | |
| zero | 35125 (48.0) | 6497 (27.8) | 27284 (60.6) | 1344 (28.4) | |
| one | 21711 (29.7) | 6645 (28.5) | 12993 (28.8) | 2073 (43.9) | <0.001 |
| two or more | 16028 (21.9) | 10207 (43.7) | 4509 (10.0) | 1312 (27.7) | |
| missing | 257 (0.4) | 0 (0) | 257 (0.6) | 0 (0) | |
| **Comorbidities** | | | | | |
| cardiovascular disease | 14597 (20.0) | 8723 (37.4) | 4657 (10.3) | 1217 (25.7) | |
| chronic obstructive pulmonary disease | 9449 (12.9) | 6467 (27.7) | 2217 (4.9) | 765 (16.2) | |
| asthma | 3552 (4.9) | 1978 (8.5) | 1464 (3.3) | 110 (2.3) | |
| diabetes | 7488 (10.2) | 5563 (23.8) | 1434 (3.2) | 491 (10.4) | |
| immunosuppression (rheumatologic/autoimmune, neoplasm) | 20288 (27.8) | 6143 (26.3) | 11779 (26.2) | 2366 (50.0) | <0.001 |
| renal disease | 3871 (5.3) | 2781 (11.9) | 895 (2.0) | 195 (4.1) | |
| cirrhosis | 1236 (1.7) | 782 (3.4) | 427 (1.0) | 27 (0.6) | |
| neuromuscular disorder | 2327 (3.2) | 1239 (5.3) | 946 (2.1) | 142 (3.0) | |
| missing | 257 (0.4) | 0 (0) | 257 (0.6) | 0 (0) | |
| **Smoking status of patient or household** | | | | | |
| currently smokes | 15872 (21.7) | 3509 (15.0) | 11644 (25.9) | 719 (15.2) | |
| has smoked previously | 13801 (18.9) | 6396 (27.4) | 6681 (14.8) | 724 (15.3) | <0.001 |
| has never smoked | 42358 (57.9) | 12634 (54.1) | 26461 (58.7) | 3263 (69.0) | |
| missing | 1090 (1.5) | 810 (3.5) | 257 (0.6) | 23 (0.5) | |

Figure S3: Distribution of all and influenza positive-only specimens by country income bracket

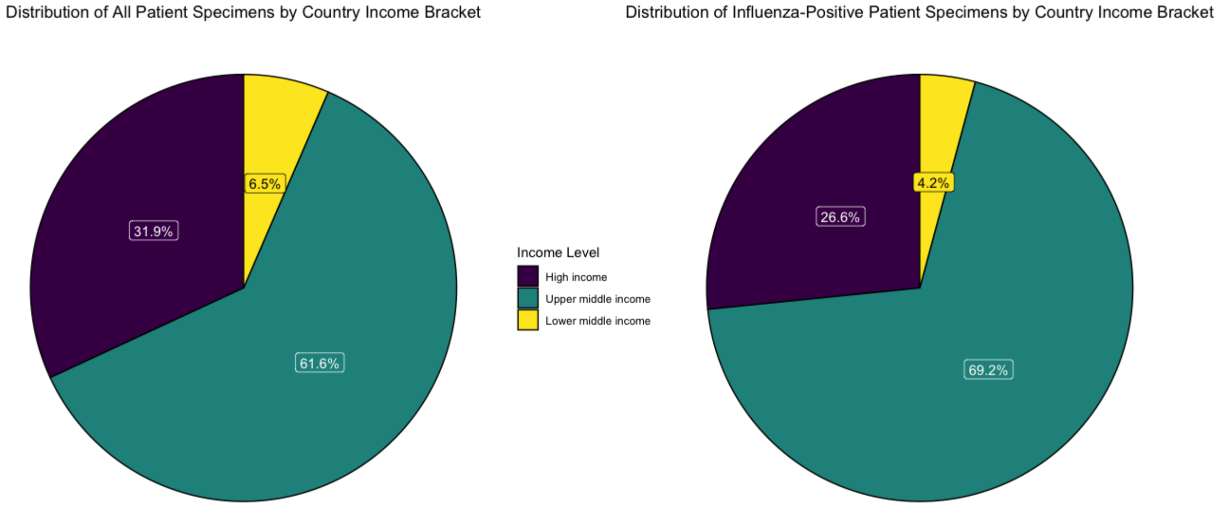

Table S4: Comparing the odds ratios of infection with A/H1N1pdm09 or A/H3N2 in patients with laboratory-confirmed influenza using Tukey contrasts

| Dataset | Outcome | Linear Hypothesis | Estimate | *p* |
|---|---|---|---|---|
| All age groups | ICU admission | A/H3N2 – A/H1N1pdm09 = 0 | -1.00 | < 0.001 |
| All age groups | In-hospital death | A/H3N2 – A/H1N1pdm09 = 0 | -0.86 | < 0.001 |
| ≥65 years only | ICU admission | A/H3N2 – A/H1N1pdm09 = 0 | -0.76 | < 0.001 |
| ≥65 years only | In-hospital death | A/H3N2 – A/H1N1pdm09 = 0 | -0.56 | < 0.05 |

Figure S4: Risk factors for (A) influenza-related intensive care unit admission, (B) influenza-related in-hospital death, (C) non-influenza intensive care unit admission, and (D) non-influenza in-hospital death, among patients from **HIC** hospitalized with severe acute respiratory illness

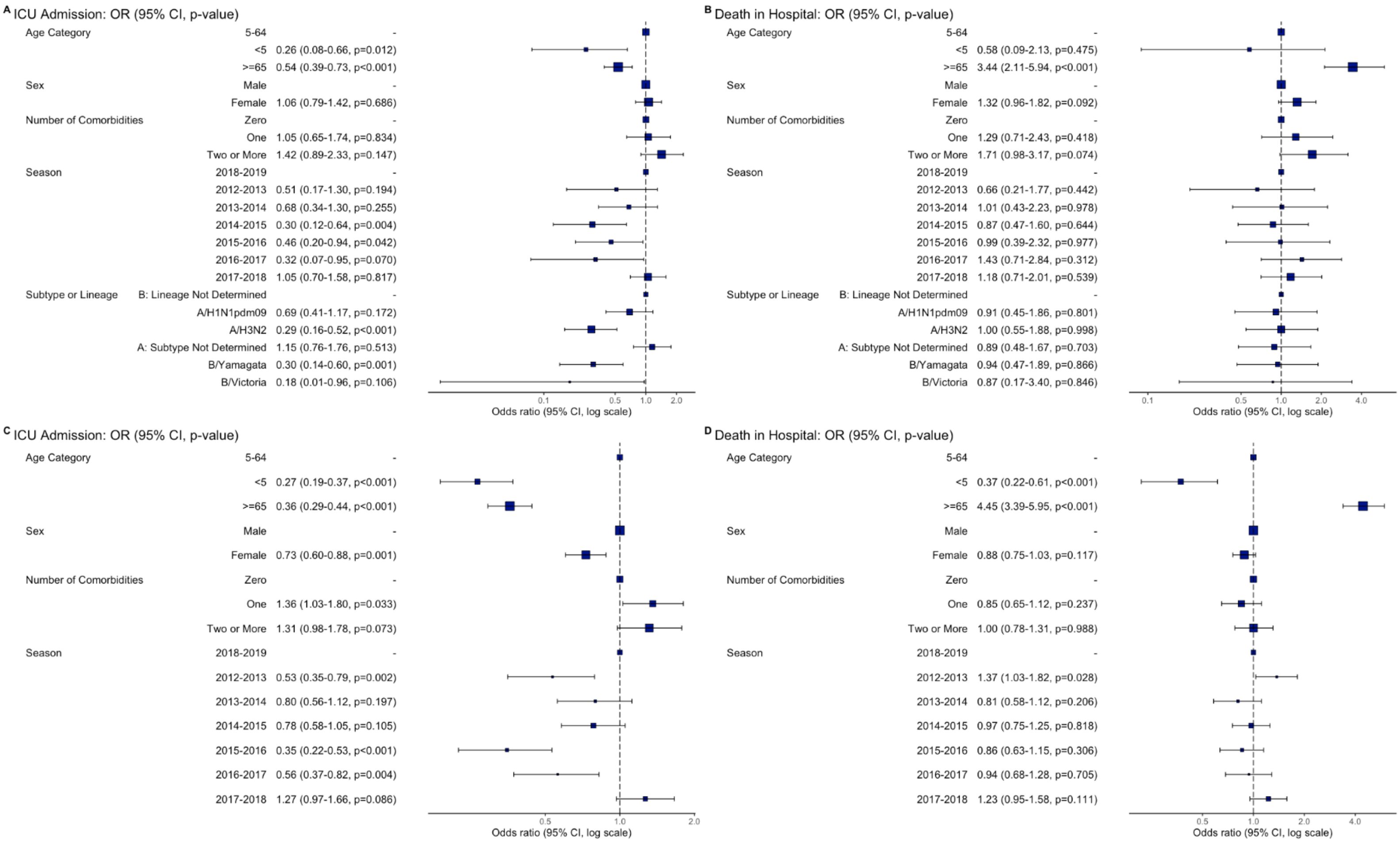
